# Application of Machine Learning (ML) to Predict Under-Five Anemia using the 2018 Zambia Demographic and Health Survey (ZDHS)

**DOI:** 10.1101/2025.08.01.25332670

**Authors:** Teebeny Zulu, Leah Kamulaza, Atupele Chisiza, Nasilele Amatende, Nathan Nasson Tembo, Mutale Sampa, Hope Sabao, Wilbroad Mutale

## Abstract

**Background:** Accurate prediction of the risk of anemia in under five children using ML can help reduce the burden of anemia in Zambia. This study applied ML models to predict the risk of anemia in under five children in Zambia.

**Methods:** This cross sectional study utilized data from the 2018 ZDHS. Feature selection was performed using the Boruta algorithm. Several ML models were trained on 80% of the data set. The best fit model was selected by comparing the model accuracy and area under the receiver operating characteristic curve (AUC -ROC). The 5-fold cross validation was used with feature importance performed using Shapley additive explanation (SHAP). Logistic regression was later performed in STATA. Python was used for ML modelling.

**Results:** Out of 7743 under five children, 58.5% had anemia. The top five most influential predictors of under-five anemia were current age of a child, maternal anemia, currently breastfeeding, region and stunting. The main ML model that was used to determine these factors was the XGB that gave an accuracy of 0.6432 and AUC-ROC of 0.6729. Two years of age or below OR=2.17, p<0.0001, currently breastfeeding OR=1.66, p<0.0001, stunting OR=1.27, p<0.0001, being a protestant OR=0.77, p=0.001, being a Muslim OR=0.33, p=0.019, living on the Copperbelt province OR=1.37, p=0.014), Luapula OR=1.91, p<0.0001), North western OR=1.66, p<0.0001, Southern OR=1.35, p=0.019, Western OR=1.47, p=0.005, having mild anemia OR=1.25, p=0.006), and moderate anemia OR=1.67, p<0.0001) significantly increased the odds of having anemia in under five children.

**Conclusion:** The extreme gradient boosting was the best performing ML model for predicting under-five mortality in Zambia and it showed that current age of a child, maternal anemia, currently breastfeeding, region and stunting are the top five most influential predictors. Therefore, this model could be useful in addressing anemia in Zambia as it can be used as a screening tool for early detection of children with anemia.

## 1. INTRODUCTION

Anemia is a significant global health concern, with children under five facing heightened vulnerability (1). Globally, an estimated 269 million children aged 6–59 months are affected by anemia (2). In Sub-Saharan Africa, the prevalence of anemia is alarming, with 67% of children affected (3,4). The repercussions of anemia in under-five children extend far beyond immediate health implications, impacting cognitive and behavioural development, physical growth, and, subsequently, affecting the socio-economic landscape of communities (2,4).

While iron deficiency remains a predominant factor contributing to anemia in various regions, the causes of this condition are intricate and context-dependent (3). Demographic characteristics, socio-economic factors, as well as maternal health factors, are also associated with the risk of anemia among children (5,6). Previous studies have found iron-deficiency anemia to be the predominant type due to diets that are low in iron and other key micronutrients such as vitamin A and folate (7–10). However, other studies have found that exclusive(11) breastfeeding beyond six months (11), younger children (6–23 months) (12), recent episodes of fever (13,14), maternal anemia (15,16), lower maternal education (17), living in poorer households (12,14,18,19) and living in households using unimproved water sources and sanitation facilities (20,21) are associated with an increased risk of anemia among under-five children. Identifying the most influential predictors of anemia in under-five children is key in developing targeted and cost-effective interventions. In addition, developing a screening tool for predicting under-five anemia based on maternal and child characteristics could supplement current diagnostic tools, especially in resource-limited settings. Machine learning models have been applied in medical research to make accurate predictions of several health conditions (6,22–24). These methods have been proven to make robust predictions compared to conventional statistical methods (25).

Despite the wide application of ML models to predict the risk of anemia in under-five children in other settings, this modelling approach has not yet been implemented in LMICs like Zambia. Many studies have identified predictors of under-five anemia in Zambia using conventional statistical approaches, which are not able to identify rank the predictors in order of their influence on anemia (10, 14, 16, 20).This study is aimed at identifying the best ML model for predicting anemia in under-five children and its risk factors so as to inform policy and ensure targeted and cost-effective interventions aimed at reducing the burden of anemia in Zambia.

## 2. Materials and Methods

### 2.1 Study Design

This study used an analytical cross-sectional study design. A detailed conceptual framework of the ML modelling steps implemented in this study was adapted from a study by Mulenga and others (27) and is presented in Figure 1

**Figure 1:**
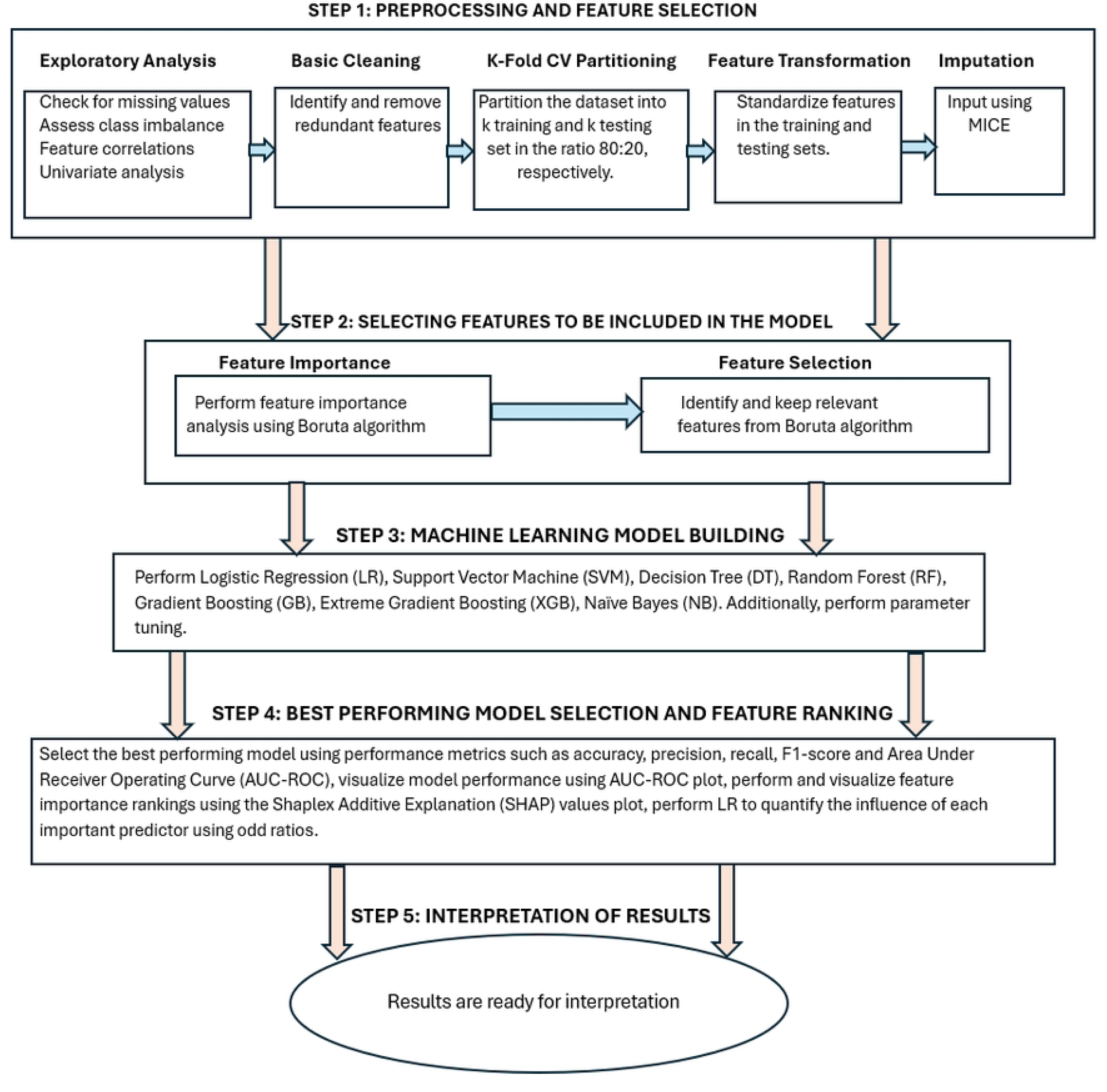
Conceptual framework of the machine learning modelling steps

### 2.2 Study population and setting

This study was conducted in Zambia which is estimated to have a population of about 21.7 million of which 14% of this population are children under the age of five (12,28). Data for this study was extracted from the children component (*kids recode*) in the 2018 ZDHS. The ZDHS is a national representative five-year periodic survey used to collect information from women aged 15–49 years and men aged 15–59 years about demographic and health status. Children aged 6-59 months from the selected household in the ZDHS sample were tested for blood hemoglobin level. In this study 7743 children aged 6-59 months with complete information of blood hemoglobin test results (anemia status) were included for analysis. Details of the DHS are published in (29).

### 2.3 Outcome variable

In this study anemia status (“*anemic” versus “not anemic”*) was the outcome variable. The original variable (*hw57*) in ZDHS was measured on an ordinal scale (*not anemic, mild, moderate and severe*). Children who had mild, moderate or severe anemia were coded 1 for *“anemic”* otherwise 0 for *“not anemic”*.

### 2.4 Predictor variables

We selected 30 variables associated with the risk for childhood anemia based on previous studies (9,14,18) and these include; maternal age, maternal education, employment status, child’s breastfeeding status, sex of a child, current age of a child, disposal of child stools when not using a toilet, type of toilet facility, source of drinking water, household wealth status, size of a child at birth, child received vitamin A in the last 6 months, distance to seek medical care at a health facility, place of antenatal care visit, under five children who slept under the mosquito bed net, food, child had fever, type of place of residence, maternal anemia, wasting, stunting, religion, region, household type of cooking fuel used, number of children ever born, number of children under five years, maternal amenorrhea, gave a child juice, number of births in the last three years and underweight.

### 2.5 Handling of Missing Data

To start with missing values in the outcome (anemia) were dropped and this reduced the sample size to 7743. Missing values in the features were imputed using multiple imputations by chained equations (MICE) in python (30).

### 2.6 Descriptive statistics and statistical software

Frequencies and percentages were presented for descriptive statistics to measure the prevalence of anemia in under-five children. Associations between under-five anemia and categorical predictors were explored using the chi-squared test and the T-test was used for quantitative predictors. Data cleaning and exploratory analysis such as checking for missing values, feature engineering, univariate analysis and LR were all performed in STATA version 17. Feature selection was performed in R version 4.4.1. The Python version 3.12.8 was using for ML model building (31).

### 2.7 Validation Strategy

This study used the 5-fold cross validation strategy where the dataset was split into the training and test sets in the ratio 80:20. This strategy has been shown to be sufficient in assessing the generalizability of ML models (35). The ML models used were optimized for performance using the GridSearchCV hyper parameter tunings (36).

### 2.8 Machine Learning Algorithms

Seven machine learning algorithms were performed in this study and these included the LR, DT, RF, SVM, GB, XGB and NB. These algorithms are discussed in detail by (6,27).

### 2.9 Performance Evaluation Metrics

The metrics used to evaluate the performance of models in this study were accuracy, precision, recall, F1-score and the AUC-ROC. The accuracy and AUC-ROC were prioritized.

### 2.10 Feature importance analysis

Feature importance was performed using the (SHAP) values after performing the XGB to rank the predictors according to their influence on anemia prediction. Logistic regression was later performed to quantify the influence that each important predictor has on anemia.

### 2.11 Ethical Approval

The ICF Institutional Review Board approved the 2018 ZDHS data survey protocols with the ICF Project Number: 132989.0.000.ZM. DHS.02. Permission to use the dataset was acquired from ICF Macro, and the dataset named ZMKR71DTA is available for download upon registration at https://www.dhsprogram.com/data. The user carefully adhered to the given guidelines, highlighting the sensitive nature of the information and the significance of not trying to uncover the identity of any household or individual surveyed for the study (maintaining anonymity).

## 3. RESULTS

### 3.1 Sample characteristics

This study involved 7743 under five children in Zambia of whom 58.5% had anemia (Table 1). Majority of children were below two years (58.2%), underweight (36.7%) and not breastfeeding (54.5%). Most of these children came from Eastern province (11.5%) with Western contributing the least (08.2%). The Mothers’ overall mean age and standard deviation was 30.0 (7.16) with most of them acquiring primary education 52.8%. Furthermore, 27.9% of these mothers had either mild, moderate or severe anemia (Table 2).

**Table 1:** Socio-demographic characteristics of a child.

| Variable | Overall<br>7743 (100%) | Anemia Status |  | p-value |
| --- | --- | --- | --- | --- |
|  |  | Not anemic<br>3217 (41.6%) | Anemic<br>4526 (58.5%) |  |
| **Current age of a child** |  |  |  |  |
| *Above 2 years* | 3236 (41.8%) | 1761(54.4%) | 1475 (45.6%) | < 0.0001<sup>CH</sup> |
| *2 years and below* | 4507 (58.2%) | 1456 (32.3%) | 3051 (67.7%) |  |
| **Currently breastfeeding** |  |  |  |  |
| *No* | 4220 (54.5%) | 1981 (46.9%) | 2239 (53.1%) | < 0.0001<sup>CH</sup> |
| *Yes* | 3523 (45.5%) | 1236 (35.1%) | 2287 (64.9%) |  |
| **Underweight** |  |  |  |  |
| *No* | 6751 (87.1%) | 2877 (42.6%) | 3874 (57.4%) | < 0.0001<sup>CH</sup> |
| *Yes* | 992 (12.8%) | 340 (34.3%) | 652 (65.7%) |  |
| **Stunting** |  |  |  |  |
| *No* | 4901 (63.3%) | 2176 (44.4%) | 2725 (55.6%) | < 0.0001<sup>CH</sup> |
| *Yes* | 2842 (36.7%) | 1041 (36.63) | 1801 ((63.4%) |  |
| **Child had fever** |  |  |  |  |
| *No* | 6376 (82.4%) | 2734 (42.9%) | 3642 (57.1%) | < 0.0001<sup>CH</sup> |
| *Yes* | 1367 (17.7%) | 483 (35.3%) | 884 (64.7%) |  |
| **Religion** |  |  |  |  |
| *Catholic* | 1267 (16.4%) | 464 (36.6%) | 803 (63.4%) | < 0.0001<sup>CH</sup> |
| *Protestant* | 6381 (82.4%) | 2708 (42.4%) | 3673 (57.6%) |  |
| *Muslim* | 31 (0.4%) | 21 (67.7%) | 10 (32.3%) |  |
| *Other* | 64 (0.8%) | 24 (37.5%) | 40 62.5%) |  |
| **Region** |  |  |  |  |
| *Central* | 762 (09.8%) | 374 (49.1%) | 388 (50.9%) | < 0.0001<sup>CH</sup> |
| *Copperbelt* | 736 (09.5%) | 315 (42.8%) | 421 (57.2%) |  |
| *Eastern* | 923 (11.9%) | 409 (44.3%) | 514 (55.7%) |  |
| *Luapula* | 891 (11.5%) | 265 (29.7%) | 626 (70.3%) |  |
| *Lusaka* | 823 (10.6%) | 368 (44.7%) | 455 (55.3%) |  |
| *Muchinga* | 708 (09.1%) | 326 (46.1%) | 382 (54.0%) |  |
| *Northern* | 800 (10.3%) | 319 (39.9%) | 481 (60.1%) |  |
| *North western* | 657 (08.5%) | 233 (35.5%) | 424 (64.5%) |  |
| *Southern* | 808 (10.4%) | 366 (45.3%) | 442 (54.7%) |  |
| *Western* | 635 (08.2%) | 242 (38.1%) | 393 (61.9%) |  |

*CH=chi-square*

**Table 2:** socio-demographic characteristics of the mother.

| Variable | Overall<br>7743(100%) | Anemia Status |  | p-value |
| --- | --- | --- | --- | --- |
|  |  | Not anemic<br>(n=6705) | Anemic<br>(n=2589) |  |
| Antenatal care visit attended |  |  |  |  |
| Elsewhere | 5083 (90.3%) | 1974(38.8%) | 3109 (61.2%) | 0.346<sup>CH</sup> |
| Government | 544 (9.7%) | 200 (36.8%) | 344 (63.2%) |  |
| Amenorrheic |  |  |  |  |
| No | 5763 (74.4%) | 2497 (43.3%) | 3266 (56.7%) | <<br>0.0001<sup>CH</sup> |
| Yes | 1980 (25.6%) | 720 (36.4%) | 1260 (63.6%) |  |
| Maternal age [Mean(SD)] | 28.95 (7.16) | 29.39(0.13) | 28.64 (0.11) | <0.0001<sup>TT</sup> |
| Number of births in the last three years |  |  |  |  |
| 0 | 1634 (21.1%) | 917 (56.1%) | 717 (43.9%) | <<br>0.0001<sup>CH</sup> |
| 1 | 5373 (69.4%) | 2035 (37.9%) | 3338 (62.1%) |  |
| 2-3 | 736 (9.5%) | 265 (36.0%) | 471 (64.0%) |  |
| Maternal education |  |  |  |  |
| No education | 840 (10.9%) | 297 (35.4%) | 543 (64.6%) | <<br>0.0001<sup>CT</sup> |
| Primary | 4091 (52.8%) | 1747 (42.7%) | 2344 (57.3%) |  |
| Secondary | 2512 (32.4%) | 1028 (40.9%) | 1484 (59.1%) |  |
| higher | 300 (03.9%) | 145 (48.3%) | 155 (51.7%) |  |
| Maternal anemia |  |  |  |  |
| Not anemic | 5601 (72.9%) | 2468 (44.06) | 3133 (55.9%) | <<br>0.0001<sup>CT</sup> |
| Mild | 1215 (15.8%) | 445 (36.6%) | 770 (63.4%) |  |
| Moderate | 824 (10.7%) | 262 (31.8%) | 562 (68.2%) |  |
| Severe | 41 (0.5%) | 20 (48.8%) | 21 (51.2%) |  |
| Children ever born |  |  |  |  |
| Below 5 years | 5154 (66.6%) | 2106 (40.9%) | 3048(59.1%) | 0.084<sup>CH</sup> |
| 5 years and above | 2589 (33.4%) | 1111 (42.9%) | 1478 (57.1%) |  |
| Type of fuel for cooking |  |  |  |  |
| *Electricity* | 406 (05.2%) | 181 (44.6%) | 225 (55.4%) |  |
| *Charcoal* | 2541 (32.8%) | 1051(41.4%) | 1490(58.6%) |  |
| *Wood* | 4612 (59.6%) | 1920(41.6%) | 2692 (58.4%) |  |
| *other* | 184 (02.4%) | 65 (35.3%) | 119 (64.7%) | **0.210<sup>CH</sup>** |

*CH=chi-square; CT=chi-square for trend; TT = T-test*

### 3.2 Feature Selection

Thirty features known to be associated with under-five anemia from literature were included in a Boruta algorithm (Figure 2) and 17 of these which included; ‘age of a child’, ‘place of antenatal care visit’, ‘disposal of child stools when not using the toilet’, gave a child juicy’, ‘food made from beans, peas, lentils and nuts’, births in the last three years’, ‘maternal anemia’, currently breastfeeding’, ‘region’, ‘stunting’, ‘underweight’, ‘residence’, ‘amenorrhea’, ‘type of fuel used for cooking’, religion’, maternal age’, having fever’ were classified as being important in predicting anemia. Of these, three features were removed as being redundant using the Cramer’s V Heatmap, leaving 14 features for model training.

**Figure 2:**
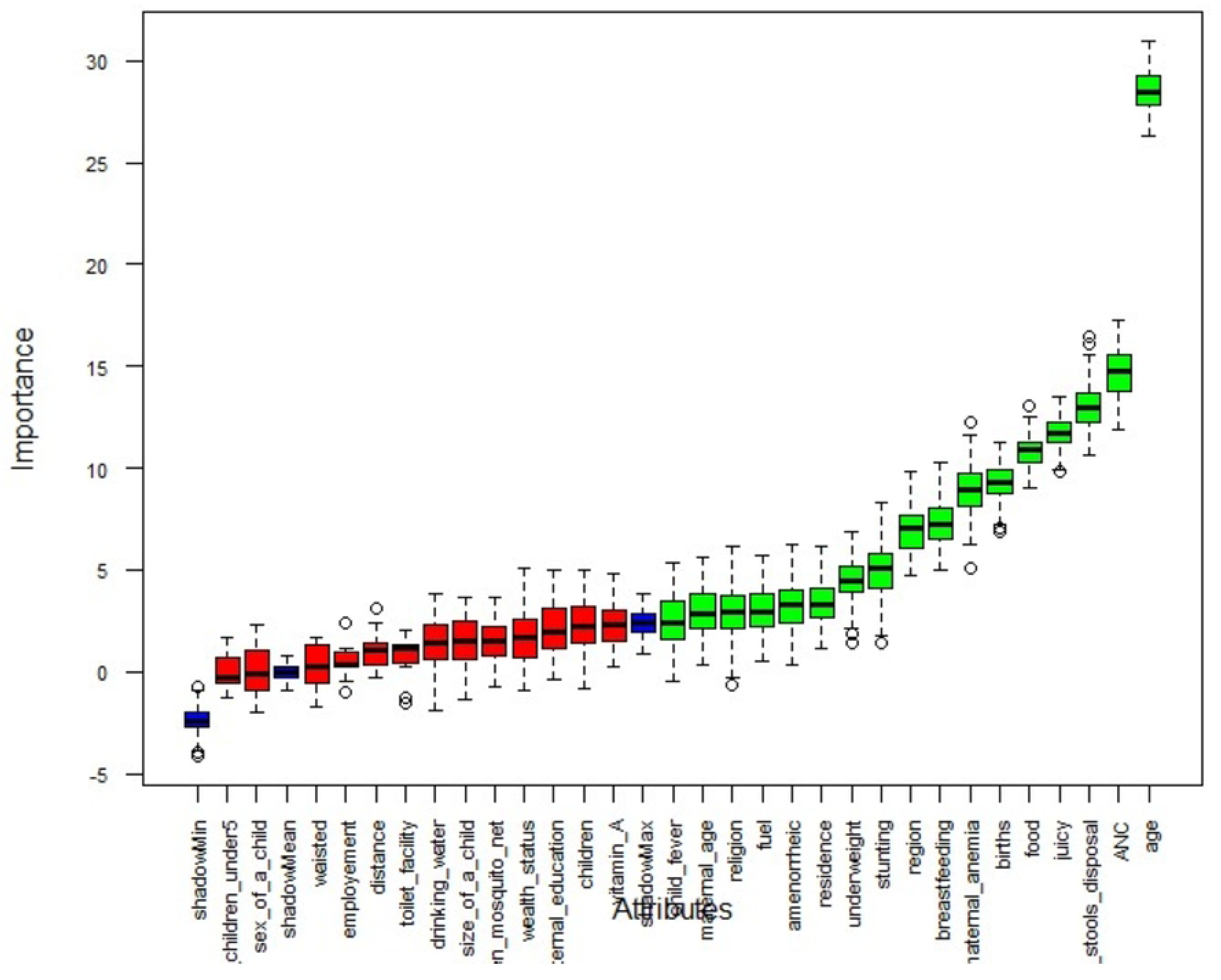
Boruta algorithm for important feature selection

### 3.3 Performance of Classification Models

Seven machine learning models were fitted in this study and the XGB model outperformed all other models with an accuracy of 0.6432 and AUC-ROC metric of 0.6729 as shown in Table 3 and Figure 3.

**Figure 3:**
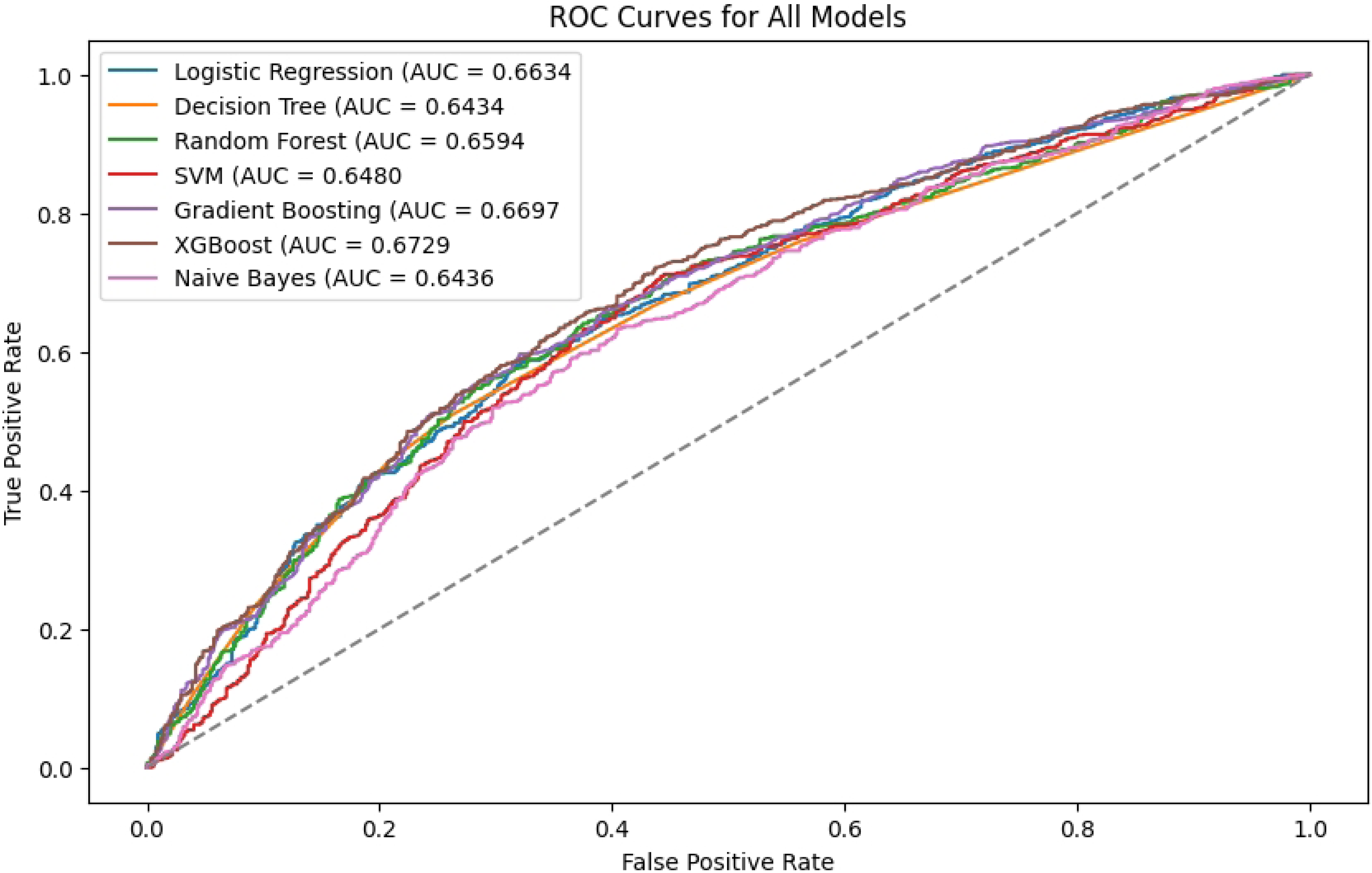
ROC curves for all ML models

**Table 3:** results from the seven ML models.

| Performance Metrics | Machine Learning Algorithms |  |  |  |  |  |  |
| --- | --- | --- | --- | --- | --- | --- | --- |
|  | LR | DT | RF | SVM | GB | XGB | NB |
| *Accuracy* | 0.6341 | 0.6208 | 0.6375 | 0.6326 | 0.6394 | 0.6432 | 0.6153 |
| *Precision* | 0.6554 | 0.6528 | 0.6595 | 0.6583 | 0.6588 | 0.6624 | 0.6717 |
| *Recall* | 0.7892 | 0.7536 | 0.7857 | 0.7726 | 0.7945 | 0.7947 | 0.6684 |
| ***F1-score*** | 0.7160 | 0.6981 | 0.7169 | 0.7106 | 0.7203 | 0.7225 | 0.6699 |
| ***AUC-ROC*** | 0.6634 | 0.6434 | 0.6594 | 0.6480 | 0.6697 | 0.6729 | 0.6436 |

*LR=Logistic Regression; DT=Decision Tree; RF=Random Forest; SVM=Support Vector Machines; GB=Gradient Boosting; XGB=Extreme Gradient Boosting; NB=Naïve Bayes.*

### 3.4 Feature Importance Analysis

Results from feature importance analysis are presented in figure 4. The first part of the plot show that the current age of a child was the most influential predictor of anemia followed by maternal anemia, currently breastfeeding, region, stunting, child fever, place of antenatal care visit, underweight, maternal amenorrhea, religion, number of births in the last three years, type of place of residence, type of fuel used for cooking, and the least was the number of children ever born. The second part of the plot shows that older age, high levels of maternal anemia, prolonged breastfeeding, stunting, child fever, antenatal care visits, and underweight increase the risk of anemia in under-five children.

**Figure 4:**
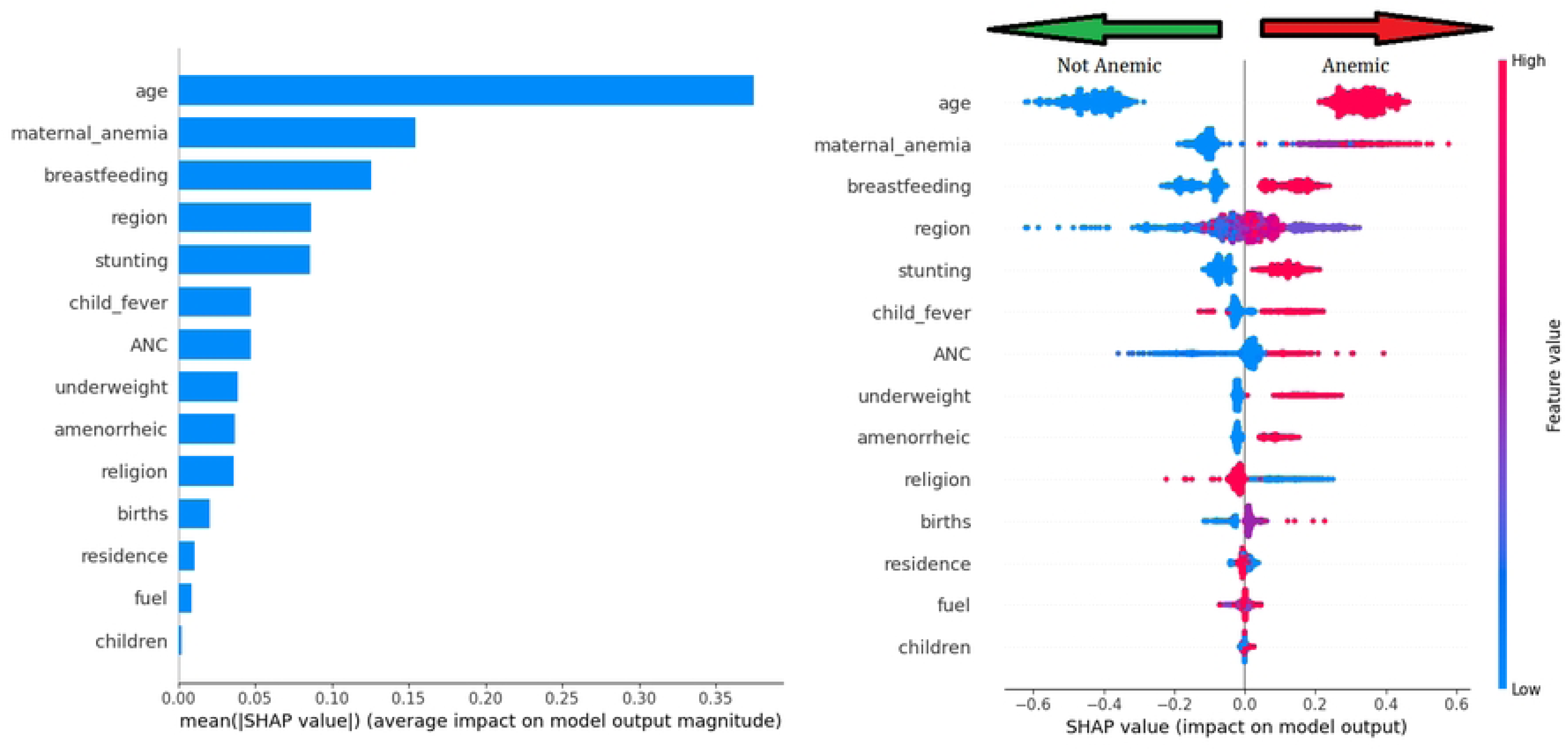
SHAP values showing feature importance analysis

### 3.5 Adjusted and unadjusted odds ratios for important predictors of under-five anemia

Results from multiple logistic regression have been grouped into child related factors (Table 4) and maternal related factors (Table 5). In this study, two years of age or below OR=2.17, (95% CI=1.6 3.05; p<0.0001), currently breastfeeding OR=1.66, (95% CI=1.44 1.92; p<0.0001), stunting OR=1.27, (95% CI=1.12 1.44; p<0.0001), being a protestant OR=0.77, (95% CI=0.65 0.90; p=0.001), being a Muslim OR=0.33, (95% CI=0.13 0.83; p=0.019), living on the Copperbelt province OR=1.37, (95% CI=1.07 1.77; p=0.014), Luapula OR=1.91, (95% CI=1.47 2.49; p<0.0001), North western OR=1.66, (95% CI=1.27 2.16; p<0.0001), Southern OR=1.35, (95% CI=1.05 1.72; p=0.019), Western OR=1.47, (95% CI=1.12 1.93; p=0.005), mild maternal anemia OR=1.25, (95% CI=1.07 1.47; p=0.006), and moderate maternal anemia OR=1.67, (95% CI=1.38 2.02; p<0.0001) significantly increased the odds of having anemia in under five children. The rest of the variables were not statically significant.

**Table 4:** Child related important predictors of anemia.

| Characteristic | UOR (95% CI) | p-value | AOR (95% CI) | p-value |
| --- | --- | --- | --- | --- |
| **Current age of a child** |  |  |  |  |
| *Above 2 years* |  |  | Ref |  |
| *2 years and below* | 2.50 (2.28 2.75) | **<0.0001** | 2.17 (1.46 3.05) | **<0.0001** |
| **Currently breastfeeding** |  |  |  |  |
| *No* | Ref |  | Ref |  |
| *Yes* | 1.64 (1.49 1.79) | **<0.0001** | 1.66 (1.44 1.92) | **<0.0001** |
| **Stunting** |  |  |  |  |
| *No* | Ref |  | Ref |  |
| *Yes* | 1.38 (1.26 1.52) | **<0.0001** | 1.27 (1.12 1.44) | **<0.0001** |
| **Underweight** |  |  |  |  |
| *No* | Ref |  | Ref |  |
| *Yes* | 1.42 (1.24 1.64) | **<0.0001** | 1.13 (0.93 1.36) | 0.216 |
| **Residence** |  |  |  |  |
| *Urban* | Ref |  | Ref |  |
| *Rural* | 1.03 (0.93 1.1) | 0.548 | 0.92 (0.78 1.09) | 0.336 |
| **Religion** |  |  |  |  |
| *Christian* | Ref |  | Ref |  |
| *Protestant* | 0.78 (0.69 .89) | **<0.0001** | 0.77 (0.65 .90) | **0.001** |
| *Muslim* | 0.28 (0.13 0.59) | **0.001** | 0.33 (0.13 0.83) | **0.019** |
| *Other* | 0.96 (0.57 1.62) | 0.887 | 1.37 (0.69 2.73) | 0.373 |
| **Child fever** |  |  |  |  |
| *No* | Ref |  | Ref |  |
| *Yes* | 1.37 (1.22 1.55) | **<0.0001** | 1.11 (0.96 1.29) | 0.153 |
| **Region** |  |  |  |  |
| *Central* | Ref |  | Ref |  |
| *Copperbelt* | 1.29 (1.05 1.58) | **0.015** | 1.37 (1.07 1.77) | **0.014** |
| *Eastern* | 1.21 (1.00 1.47) | **0.051** | 1.13 (0.89 1.43) | 0.335 |
| *Luapula* | 2.28 (1.86 2.79) | **<0.0001** | 1.91 (1.47 2.49) | **<0.0001** |
| *Lusaka* | 1.19 (0.98 1.45) | 0.082 | 1.25 (0.98 1.60) | 0.078 |
| *Muchinga* | 1.13 (0.92 1.39) | 0.244 | 0.97 (0.75 1.25) | 0.821 |
| *Northern* | 1.45 (1.19 1.78) | **<0.0001** | 1.13 (0.87 1.46) | 0.350 |
| *North western* | 1.75 (1.42 2.17) | **<0.0001** | 1.66 (1.27 2.16) | **<0.0001** |
| *Southern* | 1.16 (0.95 1.42) | 0.133 | 1.35 (1.05 1.72) | **0.019** |
| *Western* | 1.57 (1.26 1.94) | **<0.0001** | 1.47 (1.12 1.93) | **0.005** |

UOR=Unadjusted odds ratio; AOR=adjusted odds ratio; CI=confidence interval; ref=reference category

**Table 5:**
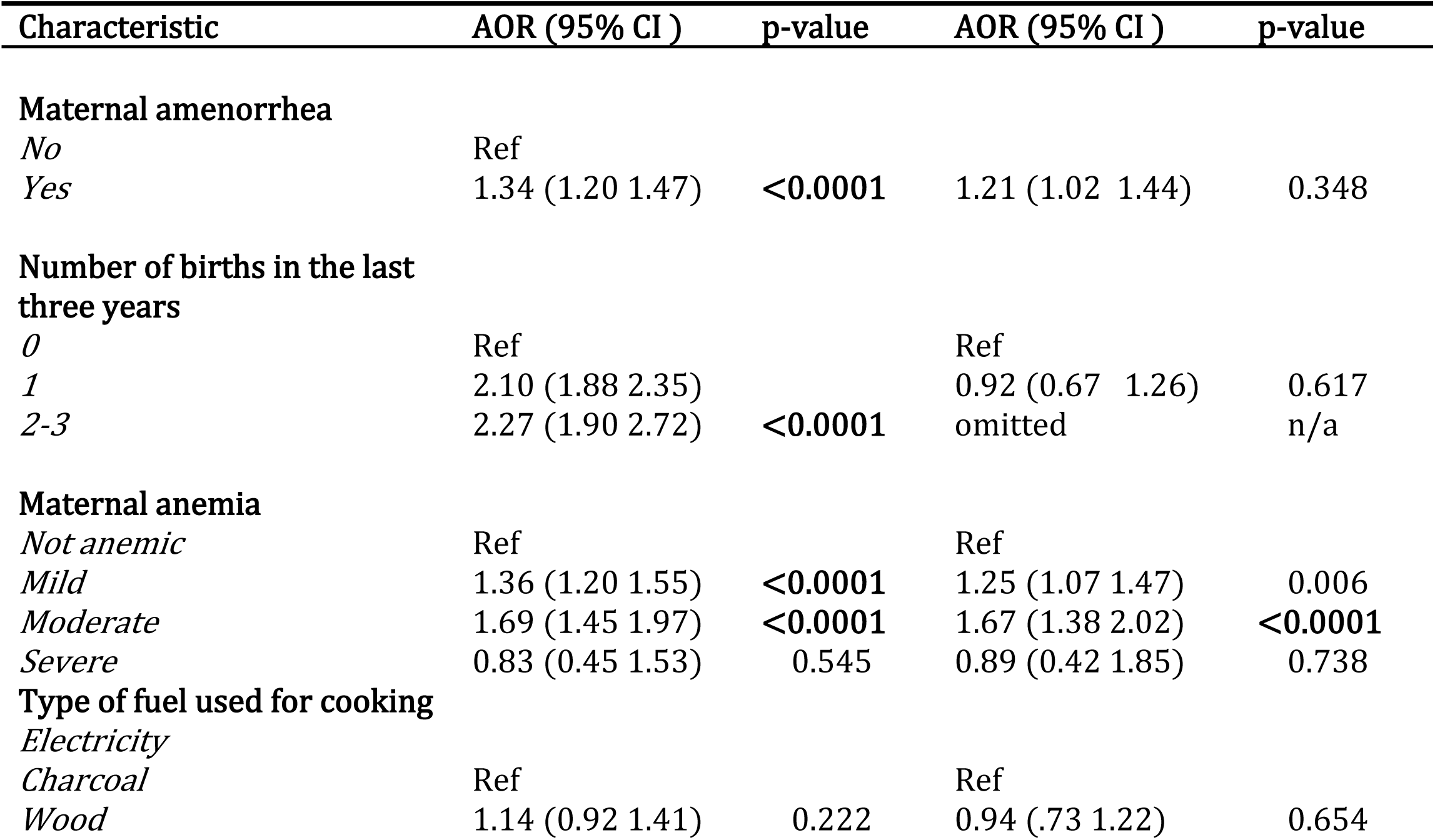
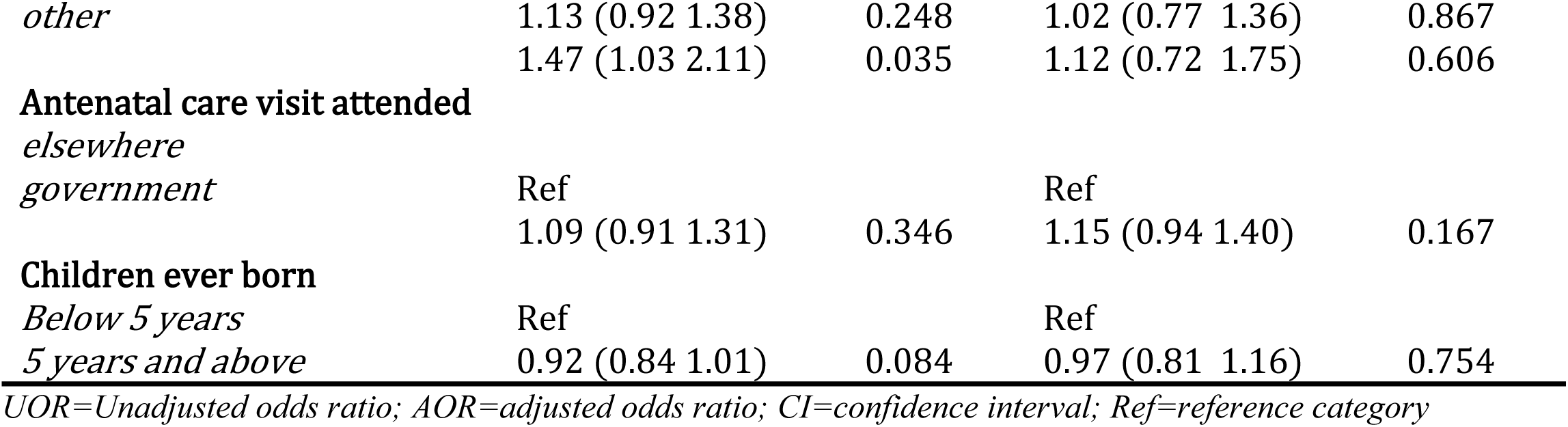
Maternal related important predictors of anemia.

## 4. DISCUSSION

### 4.1 Machine learning model performance

This study found XGB to be the best performing model with an accuracy of 0.6432 and a ROC-AUC of 0.6729. The performance of the XGB model in our study was slightly lower than the performance of a RF model in a study conducted in Ethiopia using the EDHS which had an ROC-AUC of 0.69 (22). Another study conducted in Bangladesh using the BDHS also found RF to be the best performing model with an ROC-AUC of 0.6857 (6). The variation in findings may be attributed to differences in data characteristics, such as sample size, population structure, anemia prevalence, and health system differences between countries. Additionally, models like XGB may perform better in datasets with complex feature interactions and less noise, as was potentially the case with the Zambian data (38).

### 4.2 Important predictors of anemia in under five children

The current age of a child was the most influential predictor of anemia in under-five children. Under-five children who were below two years of age were more likely to have anemia compared to those above two years. This relationship is due to elevated iron requirements because of increased growth velocity during this period which depletes iron stores acquired during pregnancy, and without appropriate iron-rich complementary foods, breast milk alone cannot meet this demand thereby increasing their susceptibility to iron deficiency. These factors highlight the importance of early interventions through iron supplementation and proper complementary feeding to prevent anemia in young children (9,39).

Maternal anemia was the second most influential predictor of anemia in this study. Logistic regression revealed that children born from mothers with mild or moderate anemia had an increased susceptibility to anemia. This relationship has been found in other studies (7,8). Maternal anemia limits the transfer of iron to the fetus, resulting in impaired fetal iron stores, which increases the likelihood of anemia after birth, especially if iron-rich complementary foods are not introduced in a timely manner (39). These findings underscore the importance of addressing maternal anemia during pregnancy to reduce the risk of anemia in children and highlight the need for early iron supplementation and appropriate feeding practices after birth.

Furthermore, breastfeeding was the third most influential predictor of anemia. Children who were currently breastfeeding were more likely to have anemia compared to those who were not breastfeeding. As infants grow, their iron needs increase, and exclusive breastfeeding beyond 6 months without the introduction of complementary iron-rich foods may lead to iron deficiency anemia (11). These results highlight the importance of timely introduction of iron-rich complementary foods to prevent anemia in breastfeeding children.

Region was ranked fourth in the top five most influential predictors of under-five anemia. Children who resided in Copperbelt, Eastern, Luapula, Northen and North western provinces of Zambia were at higher risk of anemia compared to those in Central province. This regional disparity could be explained by several contextual factors. Provinces such as Luapula, Northern, and North Western are characterized by higher poverty rates and food insecurity, which limit access to iron-rich and diverse diets essential for preventing anemia (12).

The fifth most influential predictor of under-five anemia in Zambia was stunting. This study has revealed that stunted children were more likely to be anemic as compared to those who were not stunted. This finding can be explained by the fact that both stunting and anemia often share common underlying causes, such as chronic malnutrition and micronutrient deficiencies. Stunting is a marker of prolonged under-nutrition, which is typically associated with inadequate intake of essential nutrients, including iron, folate, and vitamin A, all of which play critical roles in hematopoiesis (2).

This study brings to light the most influential predictors of under-five anemia in Zambia which is key in developing targeted interventions aimed at these predictors thereby reducing the burden of anemia in under-five children. Moreover, the identification of XGBoost as a best performing model in predicting anemia in under-five is the initial step in deploying this model to build a screening tool for under-five anemia from maternal and children characteristics. This tool could be used in resource limited settings like Zambia reducing the cost of anemia screening.

### 4.3 Strengths and limitations

This study has a number of strengths. Firstly, the use of a nationally representative sample means that these findings could be applied to all under five children in Zambia. Secondly, the use of LR through conventional statistical modelling goes beyond ranking the predictors to quantifying and explaining the influence that each predictor has on under five anemia. Despite these strengths, the notable limitation of this study is the use of the 2018 ZDHS which was taken Seven (7) years ago and may not reflect the current trends and interactions among variables. We therefore recommend that this model should be updated with the latest information to improve its prediction accuracy.

## 5. CONCLUSION

We compared seven machine learning prediction models for predicting under-five anemia. Among the models considered, the extreme gradient boosting performed the best with the highest classification accuracy and the area under the receiver operating curve. This study goes beyond identifying the best performing machine learning model and ranking predictors from the most influential to the least, to quantifying their influence with a logistic regression from the conventional statistical modelling approach. Further research on this topic should focus on deploying the machine learning model proposed in this study to build a screening tool for under-five anemia in Zambia.

## Data Availability

Permission to use the dataset was acquired from ICF Macro, and the dataset named ZMKR71DTA is available for download upon registration at https://www.dhsprogram.com/data.

